# Plasma cytokine levels reveal deficiencies in IL-8 and gamma interferon in Long-COVID

**DOI:** 10.1101/2022.10.03.22280661

**Authors:** Elizabeth S. C. P. Williams, Thomas B. Martins, Harry R. Hill, Mayte Coiras, Kevin S. Shah, Vicente Planelles, Adam M. Spivak

## Abstract

Up to half of individuals who contract SARS-CoV-2 develop symptoms of long-COVID approximately three months after initial infection. These symptoms are highly variable, and the mechanisms inducing them are yet to be understood. We compared plasma cytokine levels from individuals with long-COVID to healthy individuals and found that those with long-COVID had 100% reductions in circulating levels of interferon gamma (IFNγ) and interleukin-8 (IL-8). Additionally, we found significant reductions in levels of IL-6, IL-2, IL-17, IL-13, and IL-4 in individuals with long-COVID. We propose immune exhaustion as the driver of long-COVID, with the complete absence of IFNγ and IL-8 preventing the lungs and other organs from healing after acute infection, and reducing the ability to fight off subsequent infections, both contributing to the myriad of symptoms suffered by those with long-COVID.

## Introduction

COVID-19, caused by a novel coronavirus known as severe acute respiratory syndrome coronavirus 2 (SARS-CoV-2), was declared a pandemic by the World Health Organization March 11^th^, 2020[1]. COVID-19 is responsible for 538.6 million infections and over 6.3 million deaths worldwide as of June 18^th^, 2022[2]. For up to half of individuals who contract the virus, acute SARS-CoV-2 infection is followed by persistent health issues [3]. These individuals suffer a myriad of symptoms that affect their daily lives, including fatigue and post-exertional malaise, respiratory and cardiac symptoms, neurological symptoms, digestive symptoms, and more (Table 1)[4, 5].

**Table 1.** Long-COVID Symptoms

| Categories: | Neurocognitive: | Respiratory: | Psychological: | Other: |
| --- | --- | --- | --- | --- |
| Symptoms: | Brain fog | General fatigue | Post-traumatic stress disorder | Ageusia |
|  | Dizziness | Dyspnea | Anxiety | Anosmia |
|  | Loss of attention | Cough | Depression | Parosmia |
|  | confusion | Throat pain | Insomnia | Skin rash |
|  | **Autonomic:** | **Gastrointestinal:** | **Musculoskeletal:** |  |
|  | Chest pain | Diarrhea | Myalgias |  |
|  | Tachycardia | Abdominal pain | Arthralgias |  |
|  | palpitations | Vomiting |  |  |

Most commonly reported symptoms associated with long-COVID[4, 5].

Several names are in use to describe this post-viral syndrome, including long-haul COVID, post-acute sequalae of SARS-CoV-2 (PASC), and long-COVID. The mechanisms driving long-COVID are still poorly understood. We defined Long-COVID syndrome patients as those who fulfilled one of the following criteria: (a) individuals whose symptoms never resolved following acute infection; (b) individuals whose COVID-19 symptoms resolved but subsequently returned; or (c) individuals who developed new symptoms approximately three months after initial infection[6, 7]. Severity of symptoms during acute infection does not appear to predispose to development of long-COVID. Both asymptomatic individuals and those hospitalized due to severe complications develop long-COVID at similar rates[3, 8, 9].

Post-viral sequelae of human coronavirus (HCOV) infections have been well documented [10–12]. Both the severe acute respiratory syndrome coronavirus (SARS-CoV) outbreak in 2003[13–17] and the Middle Eastern respiratory syndrome coronavirus (MERS-CoV) outbreak starting in 2012[18–20] caused post-viral syndromes with similar symptom profiles to those experienced by individuals with long-COVID[21]. Additionally, other human coronaviruses; especially HCOV 229E and HCOV NL63; that did not reach pandemic status, have been implicated as the etiology of Kawasaki Syndrome[10, 11]. Chikungunya Virus also induces a post-viral syndrome, which presents with symptoms reminiscent of rheumatoid arthritis[22–25].

Long-COVID symptomatology bears similarities to myalgic encephalomyelitis/ chronic fatigue syndrome (ME/CFS). ME/CFS is characterized by 6 months or more of constant or relapsing bouts of excessive fatigue, cognitive impairment, post-exertional malaise, unrefreshing sleep, headaches, and neuroendocrine and immune alterations[26–30]. The number of people affected by ME/CFS is growing each year, currently affecting 0.3%-2.5% of the population globally, depending on the diagnostic criteria used[31, 32]. The heterogeneous symptoms of ME/CFS are linked to dysregulation of multiple biological systems including the immune system and inflammation[33–37], cytokines[27, 32, 38], metabolism[34, 39–42], mitochondrial function[34, 39, 43], oxidative stress[34, 36], apoptosis[34, 36], and circadian rhythm[34, 44]. Additionally, research into cytokine levels as biomarkers for ME/CFS diagnosis or severity metrics has yielded conflicting reports, for a thorough review please see Blundell *et al*. 2015[32]. There are also disparate theories regarding the origin of ME/CSF including dysbiosis of one’s microbiome[41, 45–47], and as a post-viral syndrome following infection with Epstein Barr Virus[48–51].

Based on the pro-inflammatory basis of other post viral syndromes, we hypothesized that Long-COVID is caused by abnormal, sustained, elevated levels of pro-inflammatory cytokines present in the blood after acute SARS-CoV-2 infection has abated. To test this, we assayed plasma from 15 healthy individuals and compared it to plasma from 12 patients at the University of Utah’s Long-COVID Clinic.

## Materials and Methods

### Study Subjects

We obtained healthy donor blood samples from individuals who were recruited under University of Utah Institutional Review Board (IRB) protocol 131664. These individuals were recruited from Salt Lake City, UT and the surrounding metro area between May of 2020 and December of 2021. For the purposes of this study, we define “healthy” as individuals who were uninfected or had been infected but recovered without the sequalae of long-COVID.

In the fall of 2021, the University of Utah opened a Long-COVID registry (IRB 140978). Individuals attending University of Utah Comprehensive COVID clinic or self-identified with Long-COVID can enroll in the registry, which includes a detailed symptom and health survey and blood draw for biobanking of plasma and PBMCs at the Cellular Translational Research Core (CTRC) at the University of Utah.

### Blood and Tissue Samples

15 mL of total blood was collected by phlebotomy-certified research staff into two BD Vacutainer EDTA Additive Blood Collection Tubes. Tubes were gently inverted 8-times to mix the blood and EDTA and were then centrifuged at 150g for 20 minutes at room temperature. Blood plasma was collected following centrifugation and cryopreserved in sterile cryovials at −80ºC. Peripheral blood mononuclear cells (PBMCs) were isolated by Ficoll density gradient (Histopaque-1077, Sigma), and were cryopreserved in 1mL aliquots in 80% complete culture media (endothelial cell media), 10% fetal bovine serum (FBS), and 10% dimethyl sulfoxide (DMSO) in sterile cryovials at −80ºC.

### Determination of Cytokine Concentration in Plasma

The Luminex based (Luminex Corp, TX) multiplexed cytokine assay was performed using a modified version of our previously published method[52, 53]. Briefly, monoclonal antibodies to human IL-2, sIL-2r, IL-4, IL-6, IL-8, IL-10, TNFα (BD Biosciences, Franklin Lakes, NJ), IL-13, IL-17, IFNγ (eBioscience-ThermoFisher Scientific, Waltham, MA), and IL-1β, IL-5 (R&D Systems Minneapolis, MN), IL-12 p35/p70 (Cell Sciences, Newburyport MA), were covalently coupled to MagPlex microsphere particles (Luminex Corporation) using a 2-step carbodiimide reaction, as previously described (Staros, Wright et al. 1986). A standard curve was generated by mixing known concentrations of recombinant human cytokine receptor IL-2r (R&D Systems), and recombinant human cytokines IL-1β, IL-2, IL-4, IL-5, IL-6, IL-8, IL-10, IL-12, IL-13, IL-17, TNFα and IFNγ (R&D Systems). Biotinylated secondary antibodies were purchased from the following sources: eBioscience-ThermoFisher Scientific (IL-1β, IL-4, IL-6, TNFα, IL-12p70, IFNγ, IL-13, IL-17) and BD Biosciences (IL-2, IL-5, IL-8, IL-10, IL-2r). Performance parameters including specimen dilution/recovery, detection capability, precision, interference due to hemolysis, specimen stability, and linearity were validated following Clinical & Laboratory Standards Institute (CLSI) guidelines.

### Statistical Analysis

The individual cytokine values for the healthy and Long-COVID cohorts were analyzed for statistical significance using unpaired, two-tailed, nonparametric Mann-Whitney tests with Prism 9.0 (GraphPad Software, San Diego, CA, USA).

## Results

Our long-COVID cohort included 12 individuals, who we compared to 15 matched, healthy controls (Table 2). The cytokines assayed included IL-1β, IL-2, sIL-2R, IL-4, IL-5, IL-6, IL-8, IL-10, IL-12, IL-13, IL-17, IFNγ, and TNFα. Figure 1 shows the plasma concentration in pg/ml of each cytokine. Individuals in the long-COVID cohort have decreased levels in most cytokines tested. Most notably, individuals with long-COVID have a 100% reduction in plasma levels of interferon gamma (IFNγ) and IL-8, yielding p-values of <0.0001 and 0.0011, respectively (Figure 1).

**Table 2.** Cohort Demographics

|  | All Participants | Sex |  | Age Ranges |
| --- | --- | --- | --- | --- |
|  |  | Male | Female |  |
| **n** | 27 | 7 | 20 | 23-70 |
| **Healthy** | 15 | 7 | 8 | 27-65 |
| **Long-COVID** | 12 | -- | 12 | 23-70 |

Demographics of our healthy and long-COVID cohorts.

**Figure 1.**
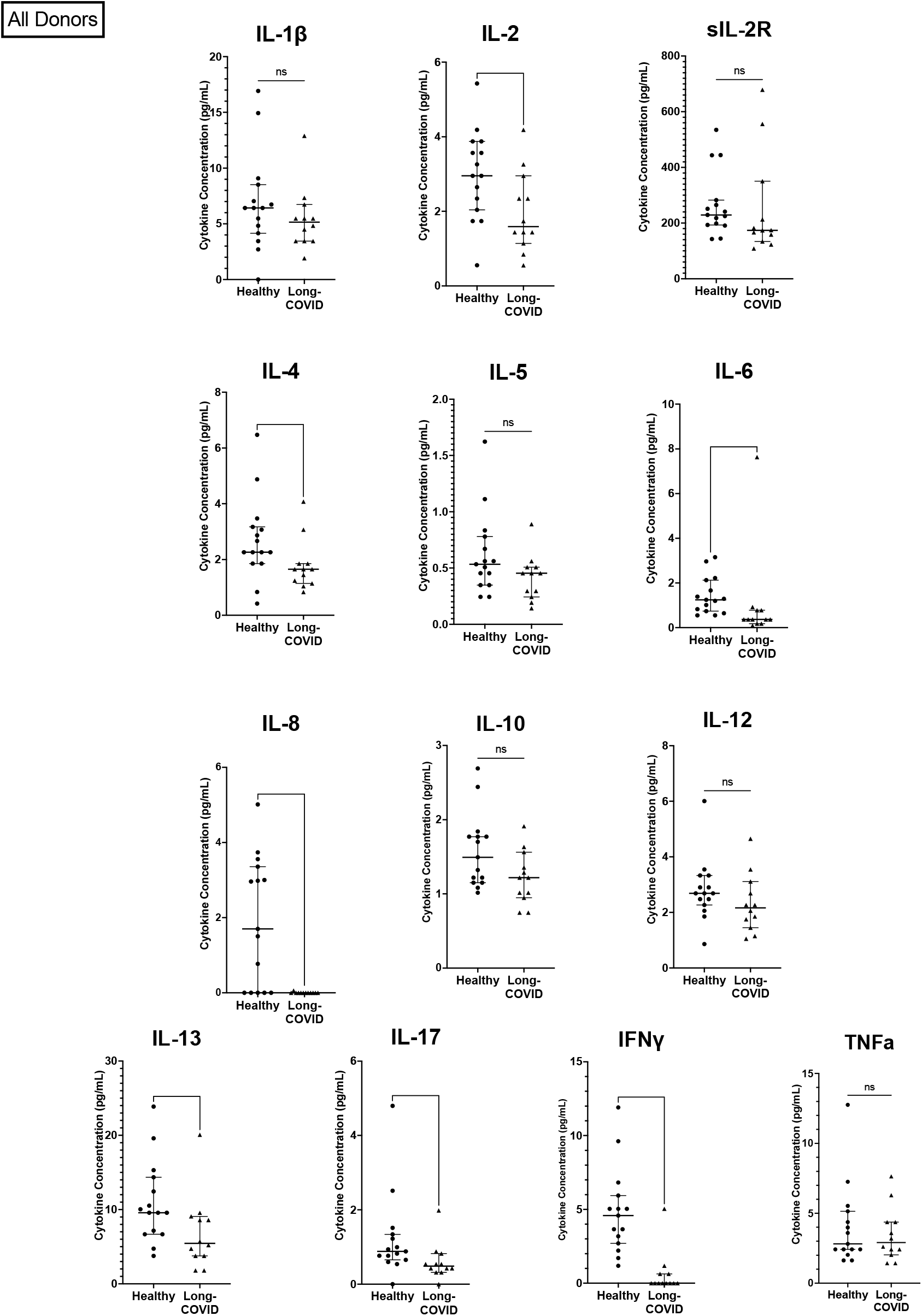
Comparison of pro-inflammatory cytokines in healthy individuals and those with Long-COVID Comparison of cytokines between the healthy cohort (n = 15) and the long-COVID cohort (n = 12). (*) p ≤ 0.05; (**) p ≤ 0.01; (***) p ≤ 0.001; (****) p ≤ 0.0001.

In addition, individuals with long-COVID have a 70% reduction in levels of IL-6. Levels of IL-2, IL-17, and IL-13 were reduced more than 40% in individuals with long-COVID (p-values 0.0285, 0.0082, and 0.0176, respectively; Figure 1; Table 3). Individuals with long-COVID also had a reduction in levels of IL-4 (26%; p = 0.0266). Differences in plasma levels of soluble IL-2 receptor (sIL-2R), IL-1β, IL-12, IL-10, IL-5, and TNFα between the long-covid and healthy groups were not statistically significant.

**Table 3.**
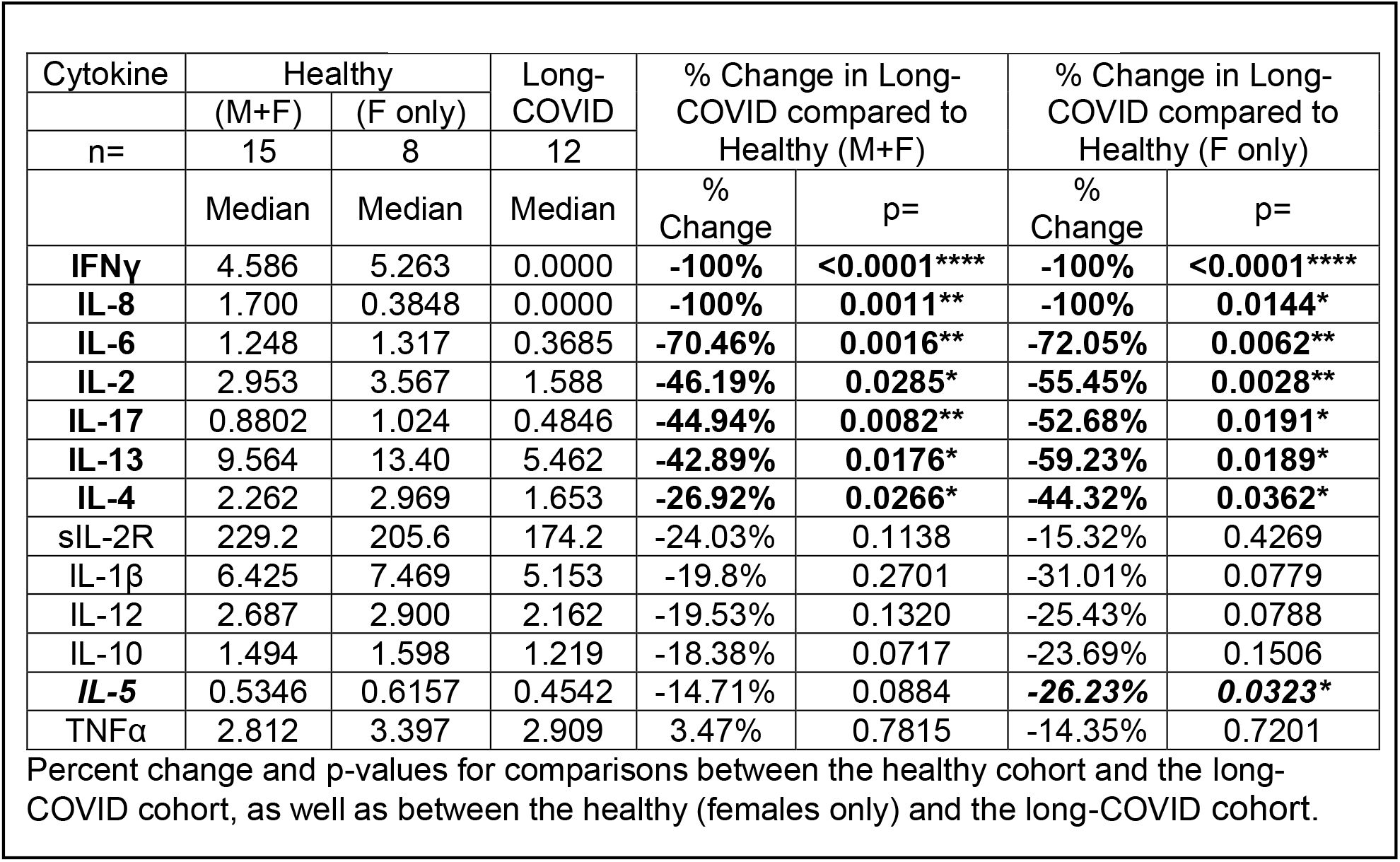
Statistical Summary of Cytokine Comparison

Given that all the participants in the long-COVID group (12 out of 12) were female (Table 2), we sought to investigate whether the observed cytokine deficits in the long-COVID group were perhaps linked to biological sex. To do this, we performed two comparisons. First, we compared cytokine levels between males and females in the healthy group. These results showed that in healthy individuals, 12 out of the 13 cytokines assayed were not significantly different between healthy males and females. One cytokine, IL-2, was 42% lower in healthy males than in healthy females (p = 0.0367; Figure 3; Table 4).

**Table 4.**
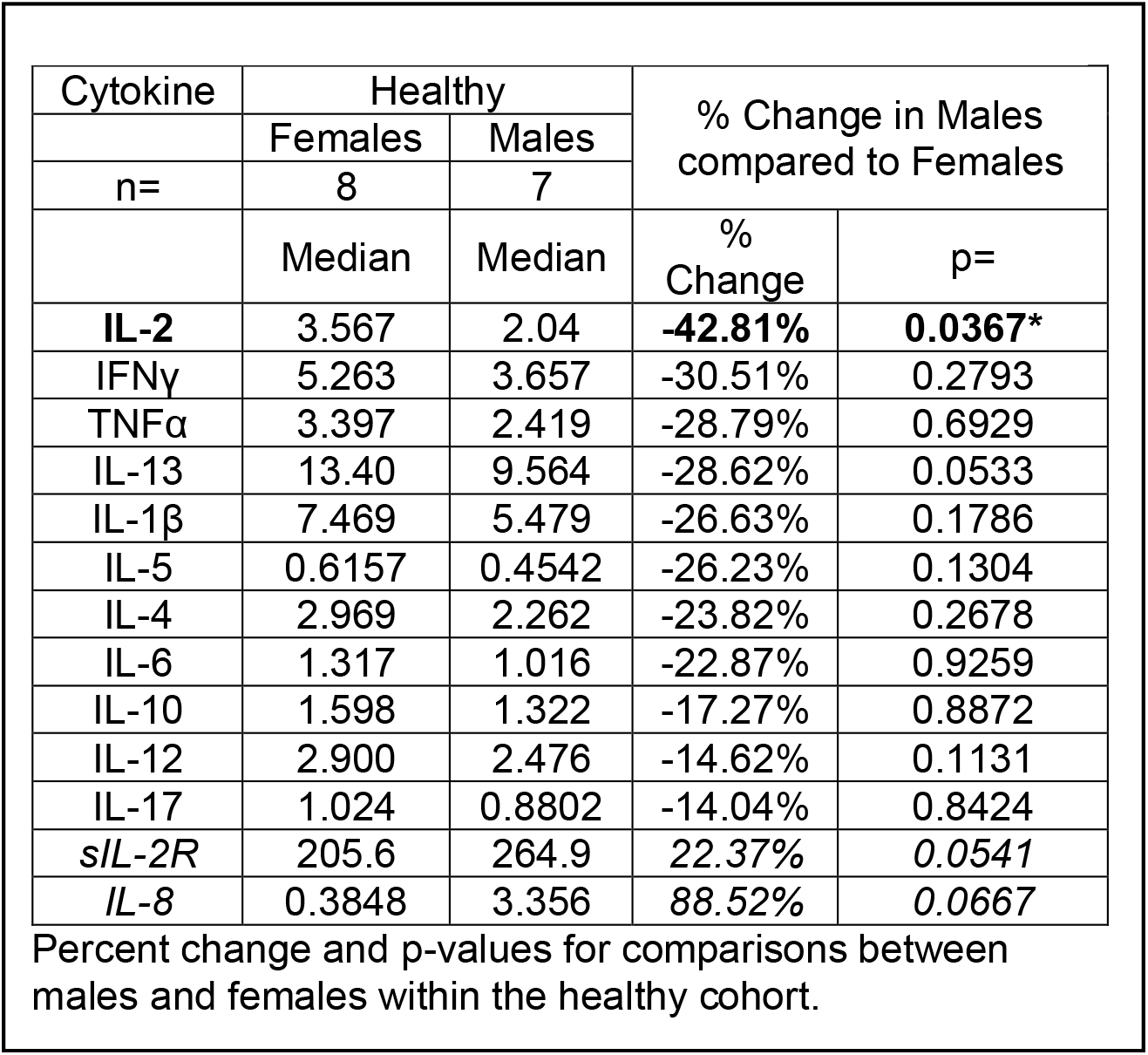
Statistical Summary of Cytokine Comparison

Secondly, we compared cytokine levels between the long-COVID group (all females) and the female participants in the healthy group (n = 8). We continue to observe a 100% reduction in IFNγ and IL-8 levels with p-values of <0.0001 and 0.0144, respectively (Figure 2). We also observed a 72% reduction in IL-6 (p = 0.0062), 55% lower levels of IL-2 (p = 0.0028), a 59% decrease in IL-13 levels (p = 0.0189), and IL-4 levels are reduced by 44% (p = 0.0362) in females with long-COVID (Figure 2; Table 3). One notable difference in the results from this female-female analysis is that the observed decrease in IL-5 levels (26%) in long-COVID females becomes statistically significant with a p-value of 0.0323, whereas the decrease between the healthy cohort when it contains both males and females and the long-COVID cohort is only 14% and is not statistically significant (Figure 1; Table 3). The changes observed in sIL-2R, IL-1β, IL-12, IL-10, and TNFα levels remain not statistically significant whether the healthy cohort includes the males or not (Figure 2; Table 3).

**Figure 2.**
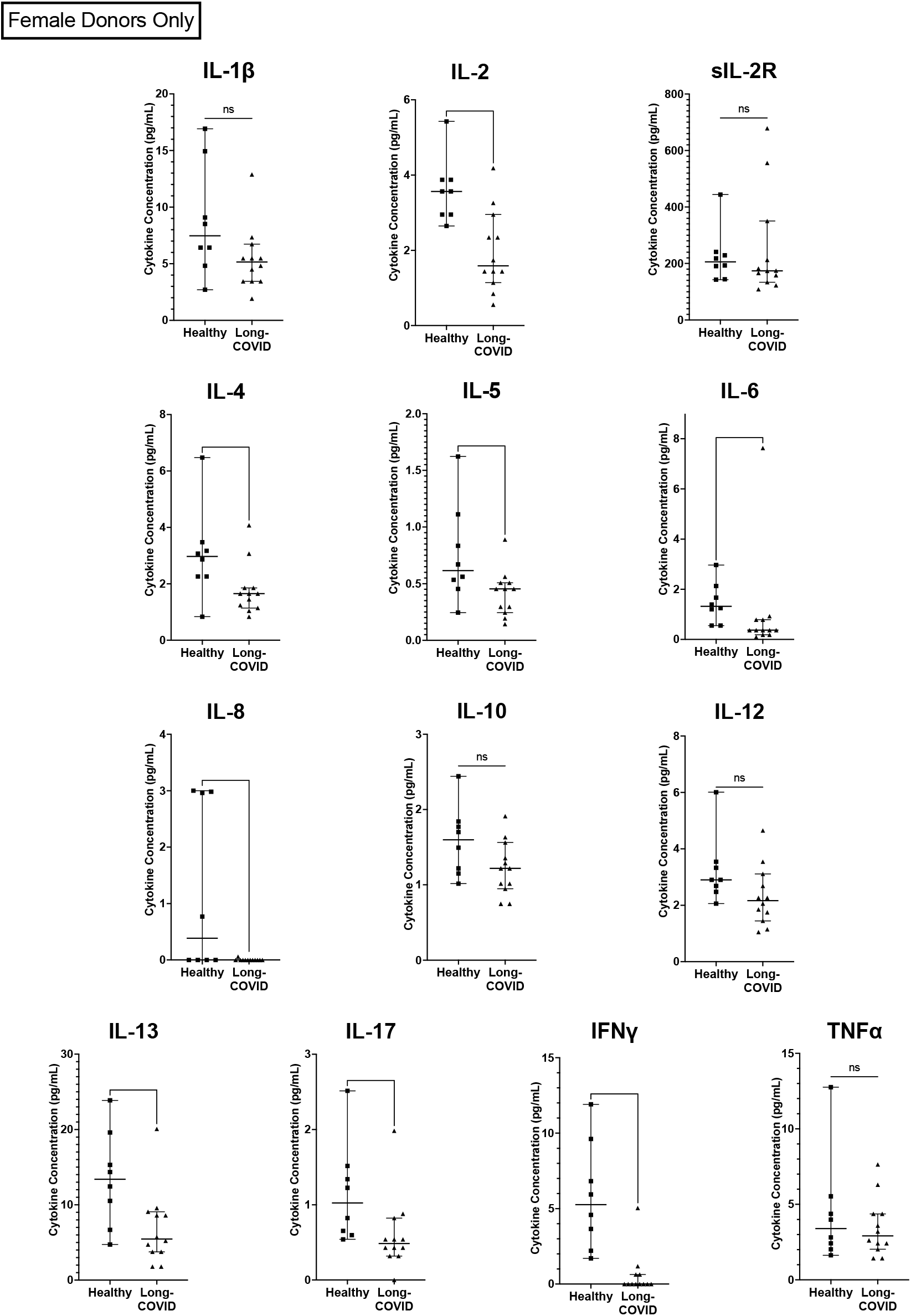
Comparison of pro-inflammatory cytokines present in healthy females and those with Long-COVID Comparison of cytokines between the healthy cohort (n = 15) and the long-COVID cohort (n = 12). (*) p ≤ 0.05; (**) p ≤ 0.01; (***) p ≤ 0.001; (****) p ≤ 0.0001.

## Discussion

Upon infection with SARS-CoV-2 the innate immune system recognizes both pathogen- and damage-associated molecular patterns (PAMPs and DAMPs, respectively) and responds by activating the NLRP3 (NOD-, LRR- and pyrin domain-containing protein 3) inflammasome[54, 55]. Monocytes and macrophages respond to PAMPs and DAMPs by secreting type I IFN and the pro-inflammatory cytokines IL-1, IL-2, IL-6, IL-12, and TNFα[54, 56]. To evaluate the possibility that dysregulated secretion of pro-inflammatory cytokines can be observed in the context of long COVID, we measured levels of 13 plasma cytokines via Luminex assay in samples from 12 donors diagnosed with long-COVID and compared them to 15 healthy controls (Table 2). All the statistically significant differences between the long-COVID cohort and healthy controls represented reductions in cytokine levels rather than the expected increases based on previous studies of other post-viral syndromes (Figure 1; Table 3)[32, 57].

Pro-inflammatory cytokines have been implicated in multiple aspects of acute COVID-19 pathogenesis. For example, increased levels of IL-1β are linked to lymphopenia in COVID-19 patients, presumably due to ongoing inflammation-induced pyroptosis[54, 58]. Macrophages express angiotensin-converting enzyme 2 (ACE2) receptors, making it possible for SARS-CoV-2 to directly infect them, and, as COVID-19 severity increases, activated macrophages congregate in the lungs, where even if not *productively* infected, an abortive infection of macrophages by SARS-CoV-2 is sufficient to induce cytokine storm[54, 59, 60]. Lastly, supporting the importance of Th17 in the pathogenesis of COVID-19, is research showing that there are increased numbers of Th17 cells present in blood samples of COVID-19 patients[54, 61].

The two most drastically decreased cytokines in our study were IFNγ and IL-8, each reduced by 100% in our long-COVID cohort. IL-8 is produced by many cell types, including epithelial cells, fibroblasts, endothelial cells, macrophages, lymphocytes and mast cells[62]. The secretion of IL-8 is induced in part by levels of IL-1β. However we found that there is no significant difference in IL-1β levels between individuals with long-COVID and healthy controls.

Also referred to as the neutrophil chemotactic factor, IL-8 recruits neutrophils and NK-cells to sites of inflammation where they can clear infected cells and promote wound healing. It is possible that the apparent lack of IL-8 in long-COVID patients may be responsible for at least some of the debilitating symptoms including post-exertional malaise, fatigue, persistent cough, shortness of breath and chest pain. In this scenario, the acute SARS-CoV-2 infection damages the lungs, the cytokine milieu unfolds as described above, recruiting cells to the site of damage where the cells can either (a) help control the infection and induce a wound healing environment and the individual recovers normally; or (b) the infection causes abundant cellular infiltration leading to a high concentration of immune cells in a relatively small physical space, ultimately causing more tissue damage, which is not efficiently repaired in the absence of IL-8.

Predictably, under scenario ‘b’ the individual remains having difficulty with oxygen transfer from the lungs into the blood stream. Therefore, if the macrophages and other cells that secrete IL-8 become exhausted or are otherwise incapable of secreting IL-8, neutrophils will not be recruited to assist in the wound healing process in the lung once the infection has been cleared[63]. Scenario ‘b’ therefore emerges as a potential model to explain certain long-COVID complications based on lack of IL-8.

IFNγ is secreted by the innate immune natural killer cells (NK) and natural killer T cells (NKT) as well as the adaptive immune CD4+ Th1 and CD8+ cytotoxic T lymphocytes (CTL) after the development of antigen-specific immunity[64]. Together with IL-12, IFNγ helps drive the differentiation of Th1 cells, which in turn can secrete IL-2, TNFα, and IFNγ [65]. The observed lack of circulating IFNγ (Figure 1; Table 2) in the plasma of COVID19 patients suggests either severe immune dysfunction or exhaustion. We observed no significant difference in the levels of IL-12 (−19%), or TNFα (3%), in individuals with long-COVID.

Levels of IL-2 and IL-4 were decreased by 46% and 26%, respectively, in individuals with long-COVID (Figure 1; Table 2). It is possible that fewer T cells differentiated into Th2 cells due to lower levels of IL-2 and IL-4, which could potentially lead to lower levels of the cytokines that Th2 cells secrete (IL-4, IL-5, IL-6, IL-9, and IL-13). This scenario may be supported by our data as we observed significantly lower levels of IL-4, IL-6, and IL-13 in individuals with long-COVID (Figure 1; Table 2). Additionally, when we compare the long-COVID cohort, which includes only females, to only the females from the healthy cohort, the decrease in IL-5 between females with long-COVID and healthy females becomes statistically significant (p = 0.0323; Figure 3; Table 2). IL-6 is involved in the differentiation of Th17 cells. It is possible that the lower levels of IL-6 we observed in long covid patients hindered the ability of the Th17 cells to properly differentiate. Supporting this possibility, we see significantly lower levels of IL-17 in individuals with long-COVID (p = 0.0082), the main cytokine secreted by Th17 cells.

**Figure 4.**
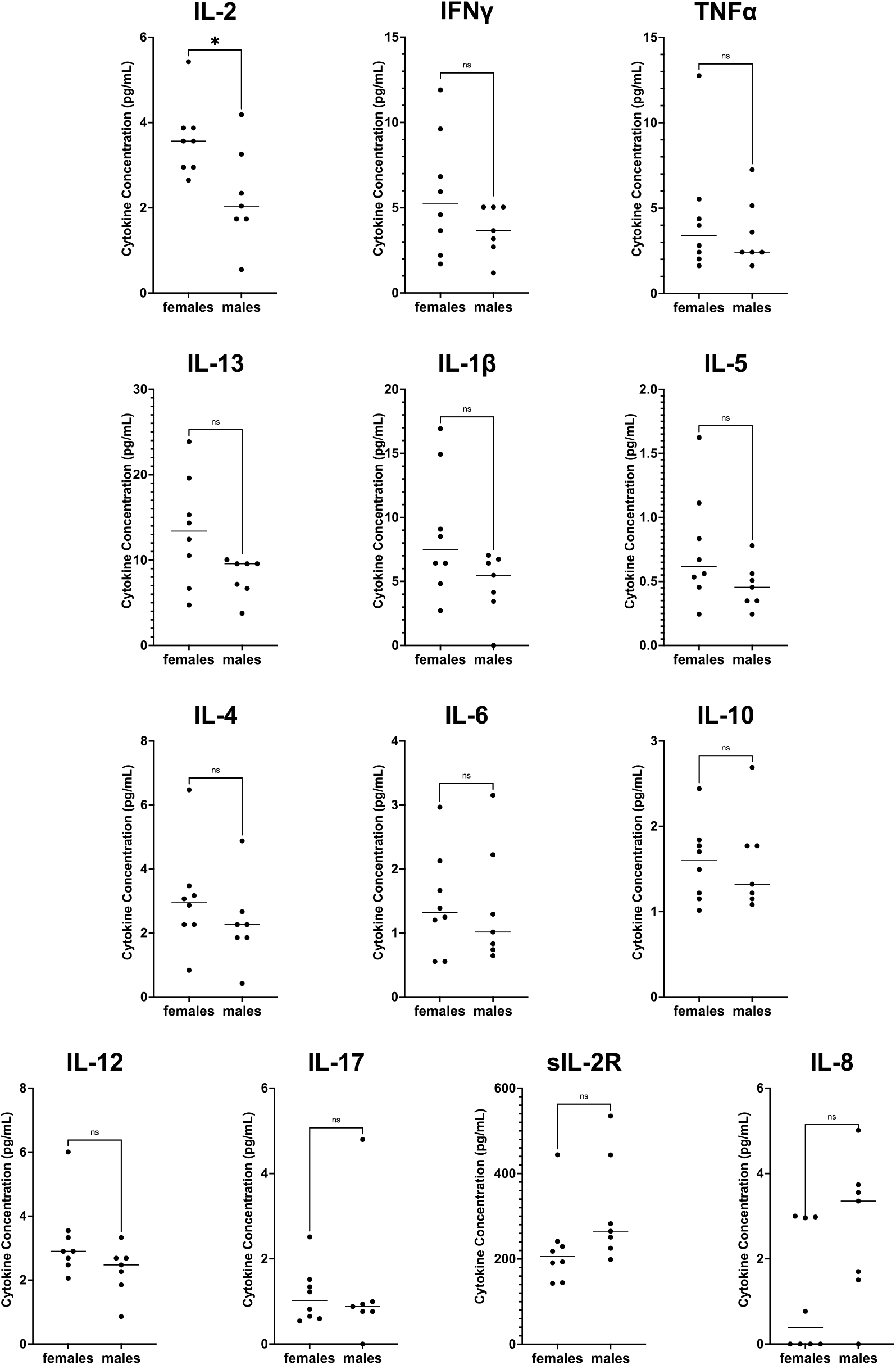
Comparison between healthy males and healthy females Compariso n of cytokines between the healthy females (n = 8) healthy males (n = 7). (*) p ≤ 0.05.

To ensure that none of the reported differences were due to inherent sex differences we compared cytokine levels between males and females within the healthy cohort. From this, we only observed one significant difference, a 42% reduction of IL-2 in healthy males compared to healthy females (Figure 3; Table 3). This sex-associated difference in IL-2 levels between healthy males and females informed us that the most accurate way to analyze IL-2 levels in our long-COVID cohort was to only consider healthy female IL-2 values. Analyzed in this way, long-COVID females show a 55% reduction in IL-2 levels (p = 0.0028; Figure 2; Table 2). The comparison including both males and females in the healthy cohort is also significant, although the inherently lower values in males complicate the interpretation.

Additionally, having re-analyzed the data to only include healthy females, it became clear that the differences that we observe between the long-COVID females, and the entire healthy population are not due solely to sex-specific differences in cytokine levels. In fact, the only cytokine that differed in having statistical significance between the healthy female to long-COVID female comparison, and the healthy male + female to long-COVID female comparison was IL-5. The removal of the male values from the analysis caused the percent change between the healthy males + females and the long-COVID females to increase from 14% to 26% (between the healthy females and long-COVID females) with a p-value of 0.0323 (Table 2; Figures 2 and 3).

Earlier we described the heterogeneous nature of the symptoms and cytokines associated with ME/CFS. Even though the symptoms of long-COVID are thought to be similar or overlapping to those of ME/CFS, when we compare our long-COVID cytokine results to those from ME/CFS patients there are glaring differences. Specifically, the pro-inflammatory cytokines IL-1β, TNFα, and IL-6 tend to be reported as being elevated in patients with ME/CFS[32], whereas we observed significant decreases in IL-6 levels in individuals with long-COVID, and no differences in levels of IL-1β and TNFα. A detailed comparison of cytokines levels published in the context of ME/CFS[32] and our long-COVID levels is provided in Supplementary Table 1.

## Conclusions

Based on our results we propose that immune exhaustion perpetuates long-COVID due to the seemingly complete reduction of IFNγ and IL-8, as well as significant decreases in IL-2, IL-4, IL-6, IL-13, and IL-17. Identifying these and other deficiencies will provide clues towards methods to intervene and possibly restore immune function in the context of long-COVID. Although functional assays that test the ability of immune cells from individuals with long-COVID to respond to pathogenic stimuli will be required to support this theory.

## Data Availability

All data produced in the present study are available upon reasonable request to the authors.

## CRediT author statement

**Elizabeth S. C. P. Williams:** Conceptualization, methodology, formal analysis, data curation, writing – original draft, visualization, funding acquisition, project administration

**Thomas Martins:** Investigation, resources

**Harry Hill:** Investigation

**Mayte Coiras:** Conceptualization, writing – review & editing

**Kevin Shah**: writing – review & editing

**Vicente Planelles:** Conceptualization, visualization, writing – review & editing, supervision, project administration

**Adam Spivak:** Conceptualization, visualization, writing – review & editing

## Ethics statements

Blood samples from donors in both our healthy and long-COVID cohorts were collected from individuals who were recruited in compliance with University of Utah Institutional Review Board (IRB) protocols 131664 and 140978, respectively.

---

**Supplemental Figure 1.**
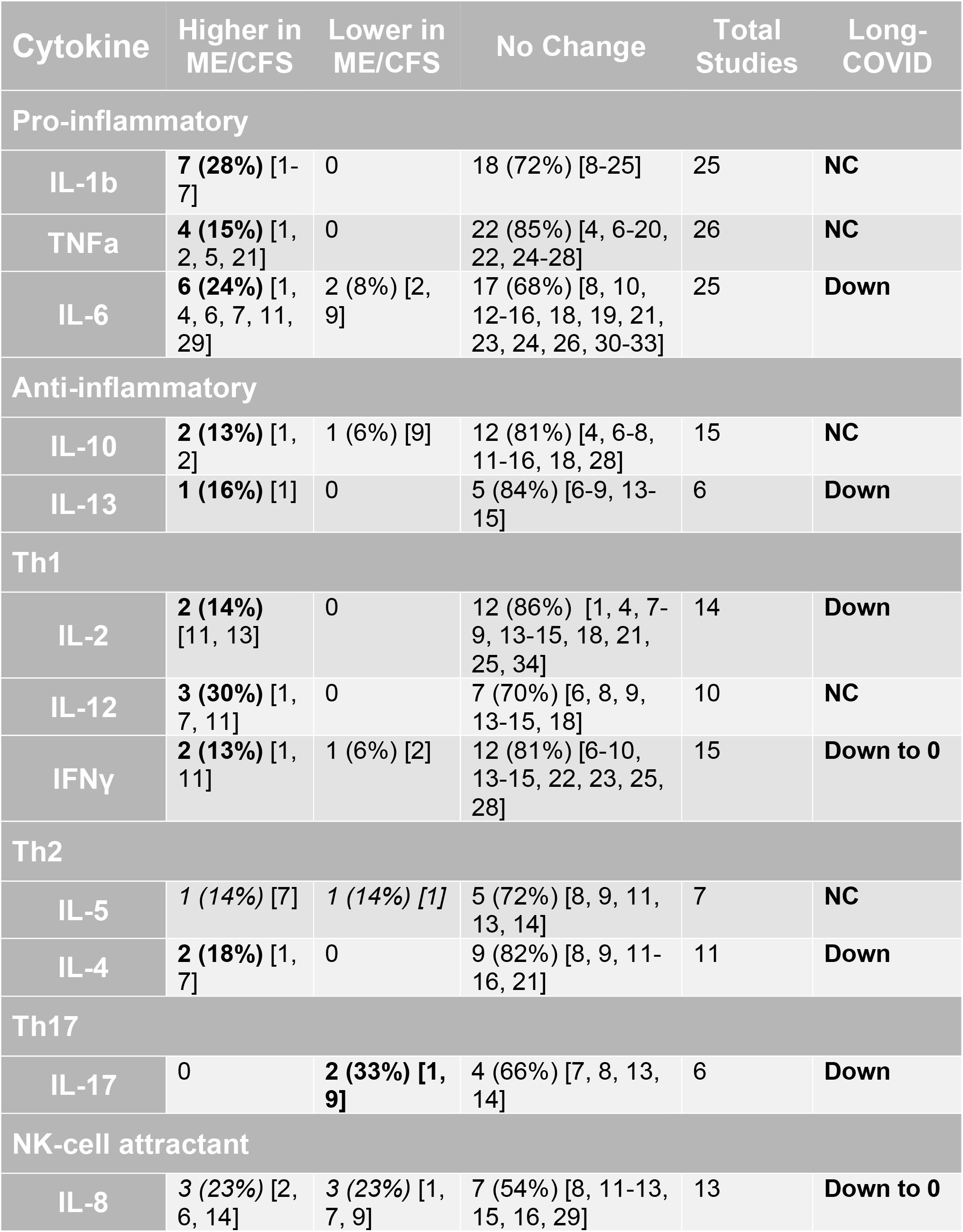
Comparison of publications depicting cytokine fluctuation in ME/CFS and our long-COVID cytokine results. Adapted from Blundell *et al*. 2015[35]. There is no agreement regarding changes in cytokine levels associated with ME/CFS. Although studies that do report changes tend to report cytokines as being upregulated in ME/CFS as compared to our long-COVID cytokine data showing significant decreases in cytokine levels.

